# COVID-19 Detection From Chest Radiographs Using Machine Learning and Convolutional Neural Networks

**DOI:** 10.1101/2020.08.31.20175828

**Authors:** Andrew C. Li, David T. Lee, Kristoff K. Misquitta, Kaiji Uno, Sasha Wald

## Abstract

Accurate and efficient diagnosis of potential COVID-19 patients is vital in the fight against the current pandemic. However, even the gold-standard COVID-19 test—reverse transcription polymerase chain reaction—suffers from a high false negative rate and a turnaround time of up to one week, preventing the infected from accessing the timely care they require, and impeding efforts to isolate positive cases. To address these shortcomings, this study develops a machine learning model based on the DenseNet-201 deep convolutional neural network, that can classify COVID-19 from chest radiographs in less than one minute and far more accurately than conventional tests (F1-score: 0.96; precision: 0.95; recall: 0.98). It uses a significantly larger dataset and more control classes than previously published models, demonstrating the promise of a machine learning approach for accurate and efficient COVID-19 screening. A live web application of the trained model can be accessed at https://cov2d19-classifier.herokuapp.com/.

## INTRODUCTION

COVID-19 is a highly infectious disease that has been declared both a Public Health Emergency and pandemic by the World Health Organization (WHO) [1]. As of August 2020, the virus has infected more than 25 million people, killed upwards of 840,000, and disrupted the lives of millions more through travel restrictions, mandated quarantine periods, and school and business closures [2].

However, these aggressive lockdowns can only be so successful when new research suggests that current screening methods are imperfect. Take, for example, the only molecular test available to diagnose current COVID-19 infection: reverse transcription polymerase chain reaction (RT-PCR) [3, 4]. Turnaround times for test results range from one day to an entire week, depending on the patient’s risk level [5]. Furthermore, within the first five days of exposure, the false negative rate for the test is 67%. Eight days after exposure, when the false negative rate is supposedly lowest, it remains as high as 20% [6]. Thus, anywhere from 20% to 67% of individuals currently screened by RT-PCR may be diagnosed as healthy when they actually have COVID-19, disrupting efforts to isolate positive cases and diverting healthcare resources from where they are most needed.

Chest radiographs (X-rays) represent another potential tool for COVID-19 diagnosis. However, some chest X-ray features of COVID-19 are only moderately characteristic to the human eye, and are sometimes undetectable [7]. Thus, diagnosis of COVID-19 by radiologists can be difficult and confusing, especially when distinguishing COVID-19 from other lung conditions like bacterial pneumonia or non-COVID-19 viral pneumonia.

Thus, an automatic COVID-19 diagnosis tool based on chest X-ray images that uses artificial intelligence and deep learning can provide two key benefits over current testing methods:

1. **Faster, cheaper, and more accurate testing**. Automated tests free up resources and streamline the healthcare process for COVID-19 patients. Machine learning has already been successfully used in the diagnosis of other medical conditions, especially cancers, and sometimes with greater accuracy than human doctors [8-9].
2. **Increased testing accessibility for burdened healthcare systems and developing countries**. Many healthcare centers already have radiological imaging machines, eliminating the logistical difficulties and expenses associated with the distribution of conventional tests. In addition, under-resourced and developing countries, where tests and human radiologists are in extremely short supply, stand to benefit from automated models that serve as alternative testing processes. In Nigeria, only 1% of the population has been tested as of August 7 due to severe test shortages, multi-week test turnaround times, and limited government laboratories. Private facilities charge $132 per test. By comparison, a chest X-ray costs less than 5 USD (1,650 Naira) in Nigeria [10].

Thus, a machine learning model based on chest radiographs has the potential to offer improvements in performance and accessibility over current methods.

In particular, convolutional neural networks (CNNs) have proven useful for automatically extracting features that are not obviously characteristic in the original image, greatly improving imaging models [11]. CNNs can be further refined using transfer learning, a technique in which the convolutional layers of a network are pre-trained on a large, unrelated dataset of images, such as ImageNet. This provides unique advantages, including faster training on the new dataset, less required data, and overall better performance.

This paper proposes a transfer-learning-based CNN to predict cases of COVID-19 from simple chest X-ray images. The model is implemented with TensorFlow and the DenseNet-201 pre-trained model trained to classify normal, bacterial pneumonia, viral pneumonia, tuberculosis, and COVID-19 pneumonia radiographs. This constitutes a total of 4 control classes apart from COVID-19.

Numerous other studies have created machine learning models to classify COVID-19 from radiological images [12-19]. However, these models are limited by their use of either a small dataset or only a few control classes. Dataset sizes in the studies reviewed range from 50 to approximately 1000 images, with only two using a dataset larger than the one in this study. Additionally, there were a maximum of only 2 control classes which always consisted of healthy images and non-COVID-19 pneumonia images. Our multiclass model uses 4 control classes: healthy, tuberculosis, and non-COVID-19 pneumonia further subdivided into two classes labelled as bacterial and viral. Combined with being trained on a larger dataset, this model has the advantage of being more generalized and robust in a real-world setting.

## METHODS

### A. Data Collection

9868 chest X-ray images of healthy individuals and those with COVID-19 pneumonia, bacterial pneumonia, viral pneumonia, or tuberculosis were compiled from several publicly available and open-source datasets [20-27]. The X-ray images were distributed as follows: 34% healthy, 28% viral pneumonia, 27% bacterial pneumonia, 5% COVID-19, and 4% tuberculosis.

**Fig. 1:**
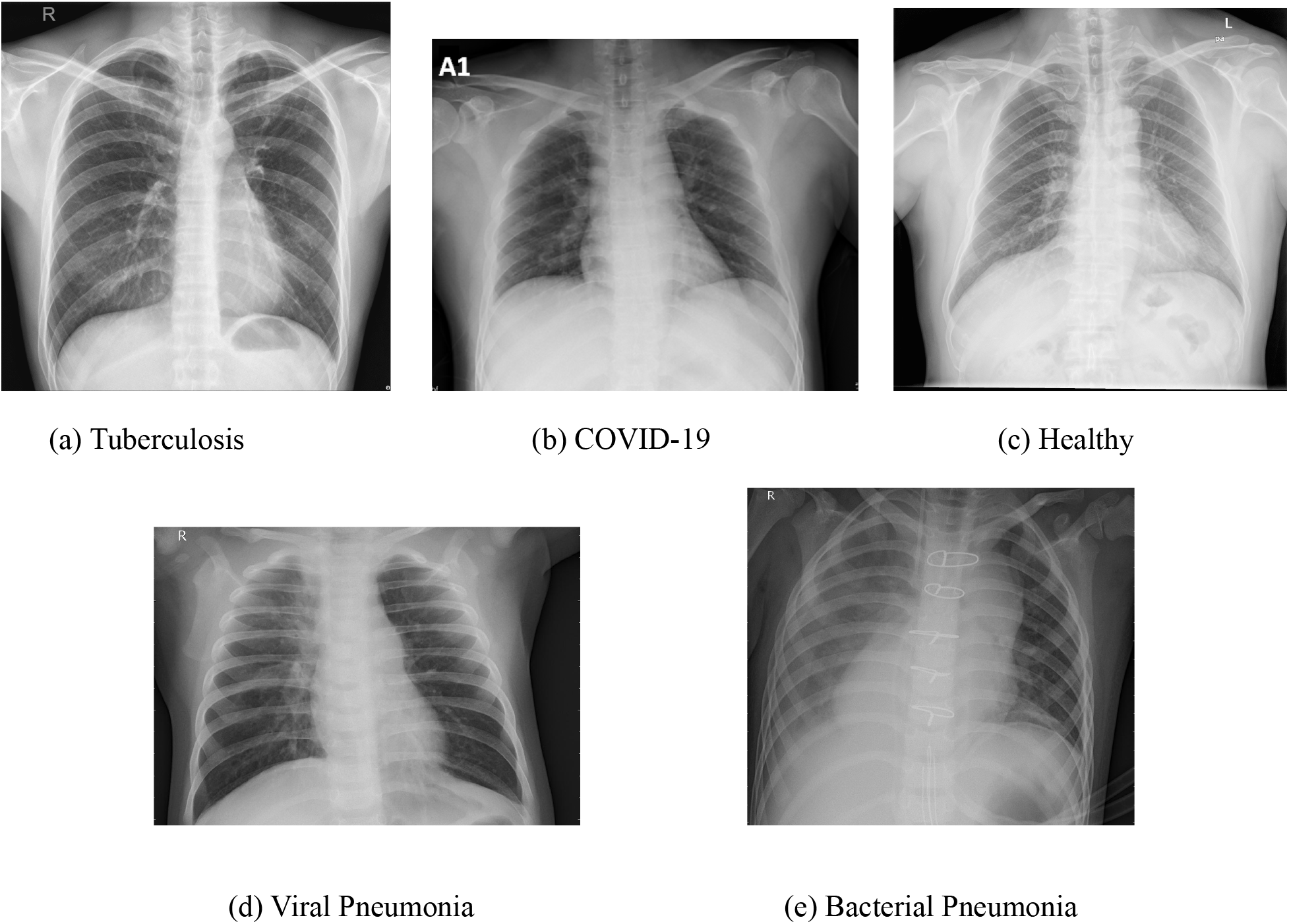
Example chest X-ray images. (a) tuberculosis, (b) COVID-19, (c) healthy, (d) viral pneumonia, (e) bacterial pneumonia

### B. Data Preprocessing

To improve model generalization and processing speed, all X-ray scans were downsampled and resized to 128×128×3 pixel dimensions. The images were then converted to RGB scale and their labels one-hot encoded to be compatible with the most common transfer learning models.

Aiming to improve model generalization and reduce overfitting, the ImageDataGenerator function of Tensorflow Keras was used to randomly augment each image in real-time before being trained on by the model. The augmentation parameters applied were horizontal flip, random rotation from 0 to 8 degrees, random width and height shift from 0% to 15%, and random zoom from 0% to 20%.

The data was then split into training, validation, and test sets. The same distribution of each disease across all split datasets was preserved. The training set consisted of 6324 images, the validation set of 1574, and the test set of 1970.

### C. Model and Architecture

DenseNet-201, trained on the ImageNet dataset, was used as the convolutional layer of the proposed model. Training was performed using the following algorithm and architecture:

1. Apply convolutions to training images using DenseNet-201, freezing the model weights.
2. Apply 2-dimensional global average pooling.
3. Flatten data.
4. Feed data into a fully-connected dense layer consisting of 32 nodes, using the ReLU activation function.
5. Feed data into a fully-connected dense layer consisting of 5 nodes, using the softmax activation function for final disease classification.

**Fig. 2:**
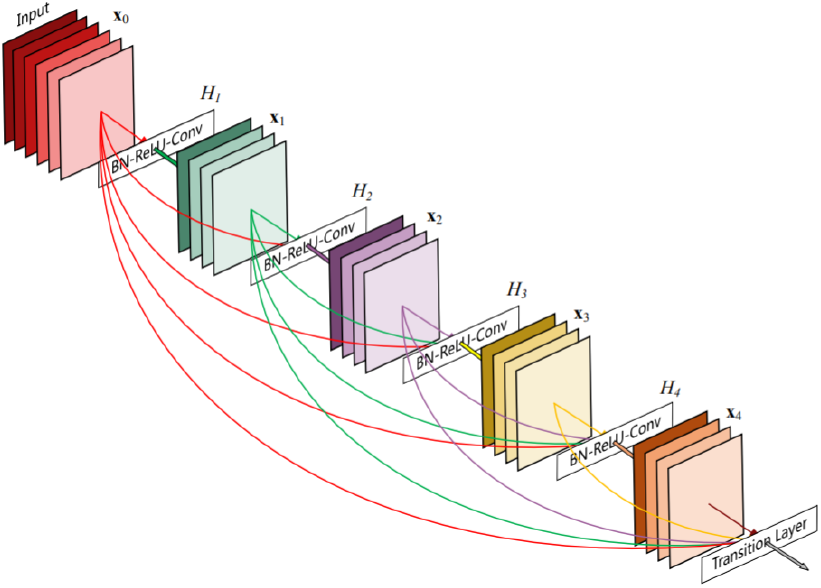
A visualization of the DenseNet-201 architecture. “Each layer takes all preceding feature-maps as input.”[28]

The default Adam optimizer and categorical cross-entropy loss function were used due to the multiclass nature of the model. Training occurred with 50 epochs and a batch size of 5. Early stopping monitoring the validation accuracy was also implemented with a patience of 5 epochs. The model was stopped at epoch 22 after validation accuracy dipped below a prior accuracy for 5 epochs in a row, indicating overfitting, and the weights of the best earlier epoch were restored for the final model.

**Fig. 3:**
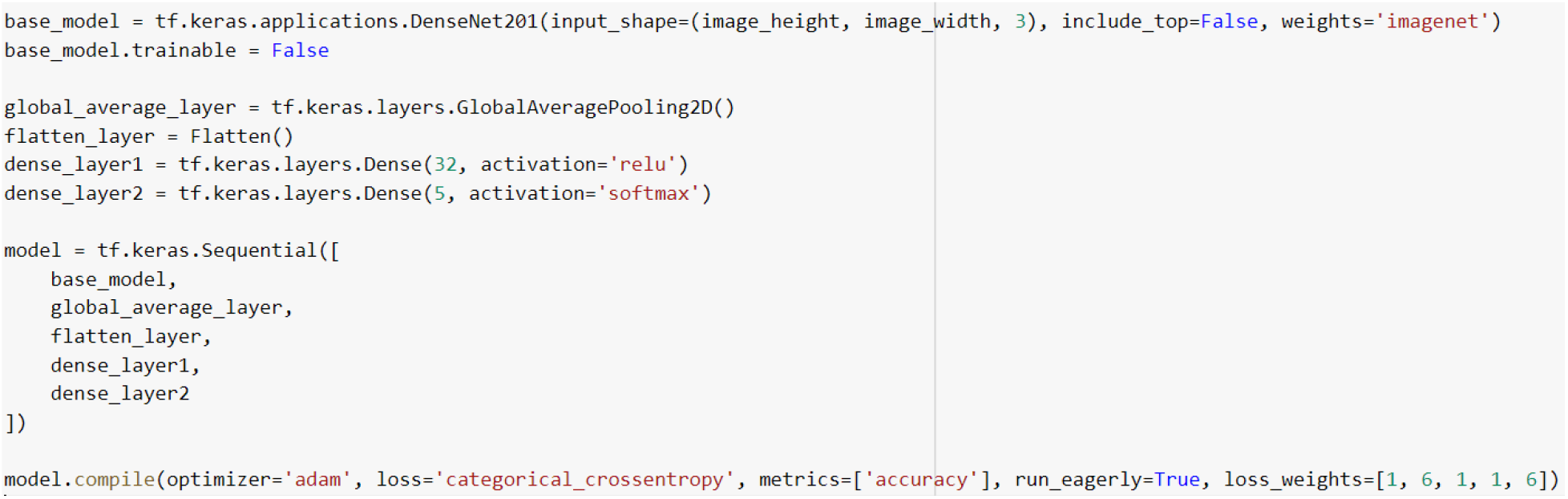
Python code used to build model with TensorFlow Keras

## RESULTS

Training accuracy steadily rose to 88.71% in the epoch which was selected for the final model. Validation accuracy stood at 82.97%, and the test set accuracy was 83.4%. More metrics are described in **Table 1**. The average area under the receiver operating curve (AUROC) for the model was 0.8963 (**Figure 4**). **Figures 5** and **6** represent a confusion matrix of the raw classification values and a heatmap of the normalized confusion matrix.

**Table 1.** Precision, recall, F1-Score, and support metrics for each class.

| Class | Precision | Recall | F1-Score | Support (Number Images in Test Set) |
| --- | --- | --- | --- | --- |
| Normal | 0.93 | 0.96 | 0.95 | 639 |
| COVID-19 | 0.95 | 0.98 | 0.96 | 125 |
| Bacterial Pneumonia | 0.76 | 0.77 | 0.76 | 556 |
| Viral Pneumonia | 0.76 | 0.74 | 0.75 | 567 |
| Tuberculosis | 0.88 | 0.73 | 0.80 | 83 |

**Fig. 4:**
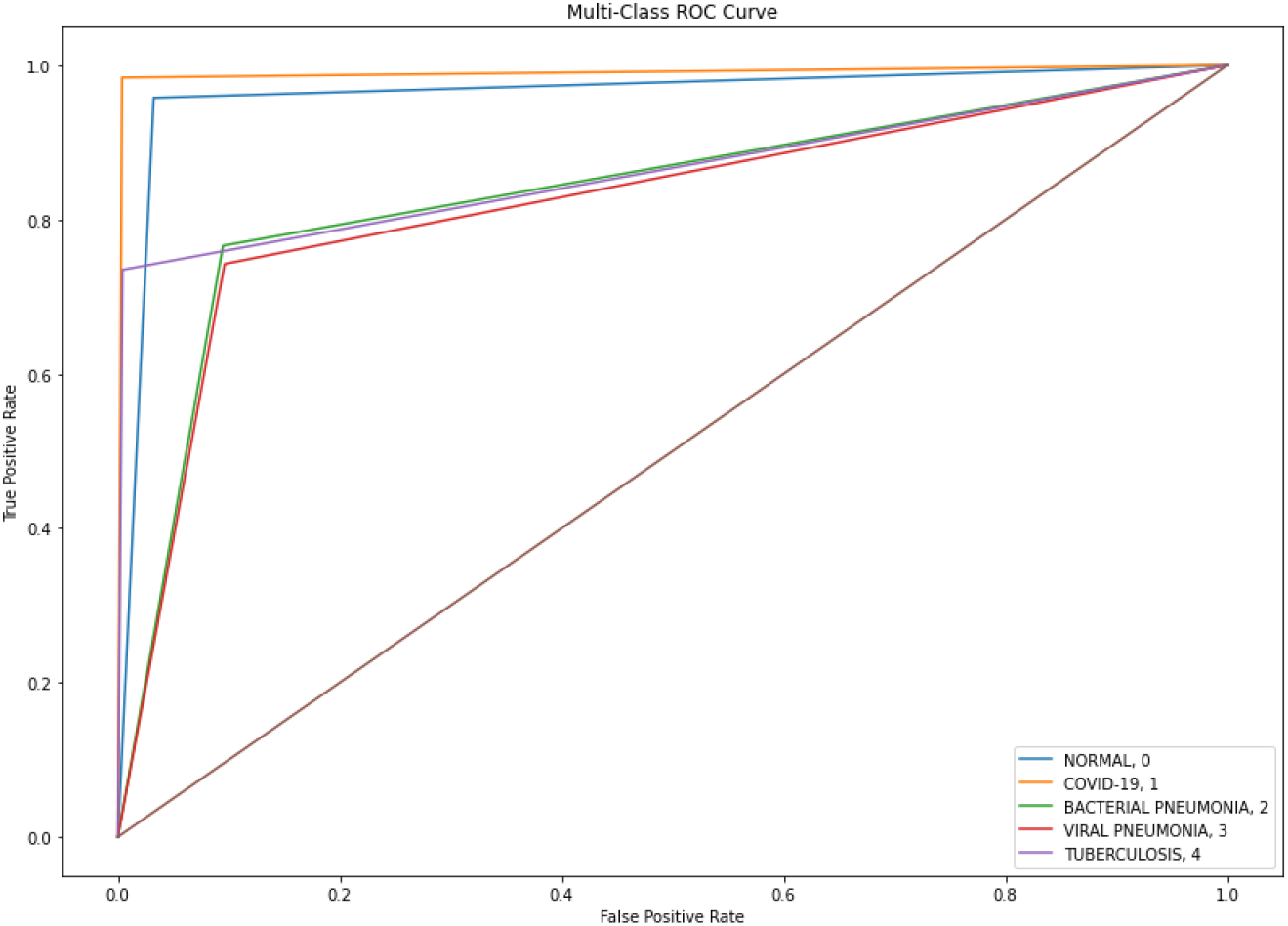
Receiver operating curve for model performance on each class.

**Fig. 5:**
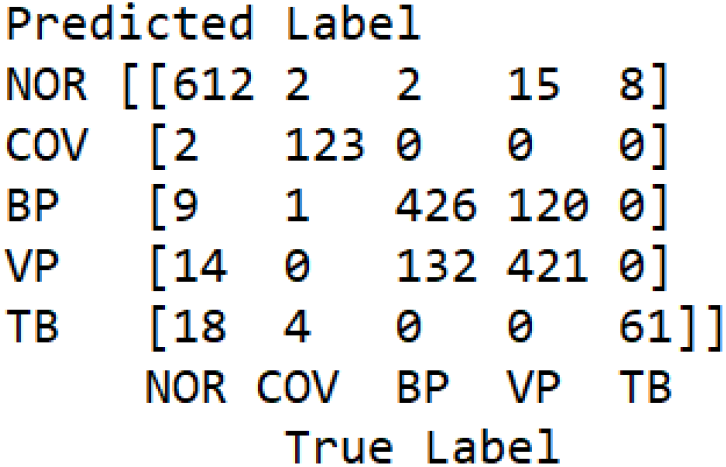
Raw confusion matrix for model test predictions. NOR: Normal, COV: COVID-19, BP: Bacterial Pneumonia, VP: Viral Pneumonia, TB: Tuberculosis.

**Fig. 6:**
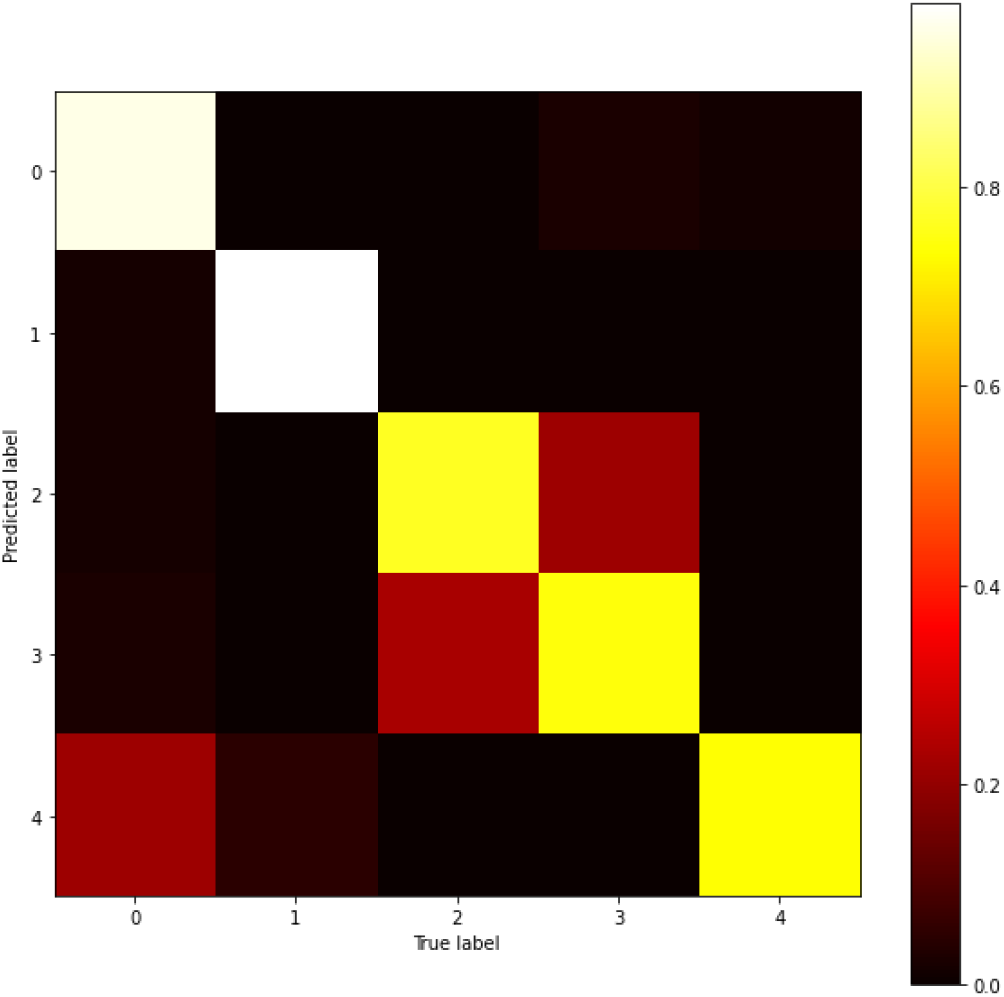
Normalized confusion matrix heatmap for model test predictions. 0: Normal, 1: COVID-19, 2: Bacterial Pneumonia, 3: Viral Pneumonia, 4: Tuberculosis.

## DISCUSSION

While our overall model classification accuracy of 83.4% could be improved upon, the classification accuracy for COVID-19 chest X-rays was exceptional, with a precision of 0.95, recall of 0.98, and F1 score of 0.96 (**Figure 4**). This represents an extremely accurate diagnosis of COVID-19: the false negative rate achieved of 5.38% is far lower than the minimum false negative rate of 20% for RT-PCR tests, and false positives are minimized as well.

Most of the reduction in the model’s accuracy can be traced to its trouble with distinguishing bacterial and viral pneumonia. While this limits the model’s usability as a general-purpose diagnostic tool across all five lung classes, it does not interfere with its primary goal of identifying new COVID-19 infections. Ultimately, the model was able to classify COVID-19 chest X-rays more accurately than common tests used today, as well as differentiate COVID-19 from numerous other lung conditions.

It should be noted that future studies should investigate using more common respiratory infections such as the common cold or seasonal flu as control classes to confirm the model’s robustness. Further, due to the contagious nature of COVID-19, the physical practicalities of using imaging equipment on COVID-19 patients are complex as the machinery must be thoroughly cleaned between each screening, and patients are at risk of transmitting the virus whilst moving to and from the machinery [29].

A live web application of the trained model can be found at https://cov2d19-classifier.herokuapp.com/. Given an X-ray image, the app will return the probability that it belongs to each class along with a final prediction. This application is in beta development stage and has not been tested in a professional environment; therefore, it is not recommended for official medical use.

## CONCLUSION

In this paper, we have developed a convolutional neural network using the DenseNet-201 model to detect cases of COVID-19 pneumonia from chest radiographs. It is robust and reliable, using a larger dataset and more control classes than most other models published. It is able to accurately distinguish COVID-19 from healthy, viral pneumonia, bacterial pneumonia, and tuberculosis X-ray images. It achieves an F1-score of 0.96 and a COVID-19 classification accuracy far exceeding that of RT-PCR tests, which suffer from a high false negative rate.

Given the sheer volume of patients that must be screened, this automated tool can save valuable time, money, and resources that are scarce in healthcare systems around the world. It may especially impact developing countries and resource-strained regions where both molecular tests and trained radiologists are in short supply, as an automated diagnosis tool provides a simple detection solution. We believe that this model, and machine learning as a whole, can significantly improve current COVID-19 diagnosis practices—vital in a pandemic where accurate and efficient screening is a must.

## Data Availability

All data is publicly available and listed in the references of the paper (references 20-27).

## ACKNOWLEDGEMENTS

This paper and model would not have been possible without the support of the MIT Lincoln Laboratory, Beaver Works Summer Institute, and Medlytics course instructors. In particular, instructors Jordan Montgomery, PhD, Jeffrey Arena, and Thomas Curtis helped immeasurably with their expertise and teaching.

## REFERENCES

[1] “Timeline of WHO’s Response to COVID-19.” World Health Organization, World Health Organization, 2020, www.who.int/news-room/detail/29-06-2020-covidtimeline.

[2] “COVID-19 Map.” Johns Hopkins Coronavirus Resource Center, 2020, coronavirus.jhu.edu/map.html.

[3] Wang, Wenling, et al. “Detection of SARS-CoV-2 in Different Types of Clinical Specimens.” Jama, 2020, doi:10.1001/jama.2020.3786.

[4] “Testing for COVID-19.” Centers for Disease Control and Prevention, Centers for Disease Control and Prevention, 24 Aug. 2020, www.cdc.gov/coronavirus/2019-ncov/symptoms-testing/testing.html.

[5] Mukherjee, Sy. “The Average Turnaround for COVID Test Results Is Now 7 Days or More.” Fortune, Fortune, 14 July 2020, fortune.com/2020/07/14/how-long-do-coronavirus-test-results-take-quest-diagnostics-covid/.

[6] Kucirka, Lauren M., et al. “Variation in False-Negative Rate of Reverse Transcriptase Polymerase Chain Reaction-Based SARS-CoV-2 Tests by Time Since Exposure.” Annals of Internal Medicine, vol. 173, no. 4, 2020, pp. 262-267., doi:10.7326/m20-1495.

[7] Ng, Ming-Yen, et al. “Imaging Profile of the COVID-19 Infection: Radiologic Findings and Literature Review.” Radiology: Cardiothoracic Imaging, vol. 2, no. 1, 2020, doi:10.1148/ryct.2020200034.

[8] Kann, Benjamin H, et al. “Artificial Intelligence in Oncology: Current Applications and Future Directions.” Cancer Network, 15 Feb. 2019, www.cancernetwork.com/view/artificial-intelligence-oncology-current-applications-and-future-directions.

[9] Walsh, Fergus. “AI ‘ Outperforms’ Doctors Diagnosing Breast Cancer.” BBC News, BBC, 2 Jan. 2020, www.bbc.com/news/health-50857759#:~:text=Artificial%20intelligence%20is%20more%20accurate,images%20from%20nearly%2029%2C000%20women.

[10] “Nigeria Healthcare Service Cost Comparison.” Knoema, 3 Aug. 2018, knoema.com/jlebqif/nigeria-healthcare-service-cost-comparison.

[11] Krizhevsky, Alex, et al. “ImageNet Classification with Deep Convolutional Neural Networks.” Communications of the ACM, vol. 60, no. 6, 2017, pp. 84-90., doi:10.1145/3065386.

[12] Apostolopoulos, Ioannis D., and Tzani A. Mpesiana. “Covid-19: Automatic Detection from X-Ray Images Utilizing Transfer Learning with Convolutional Neural Networks.” Physical and Engineering Sciences in Medicine, vol. 43, no. 2, 2020, pp. 635-640., doi:10.1007/s13246-020-00865-4.

[13] Asif, Sohaib, et al. “Classification of COVID-19 from Chest X-Ray Images Using Deep Convolutional Neural Networks.” *MedRxiv,* 18 June 2020, doi:10.1101/2020.05.01.20088211.

[14] Ozturk, Tulin, et al. “Automated Detection of COVID-19 Cases Using Deep Neural Networks with X-Ray Images.” Computers in Biology and Medicine, vol. 121, June 2020, p. 103792., doi:10.1016/j.compbiomed.2020.103792.

[15] Sethy, Prabira Kumar, and Santi Kumari Behera. “Detection of Coronavirus Disease (COVID-19) Based on Deep Features.” 2020, doi:10.20944/preprints202003.0300.v1.

[16] Xu, Xiaowei, et al. “A Deep Learning System to Screen Novel Coronavirus Disease 2019 Pneumonia.” Engineering, 27 June 2020, doi:10.1016/j.eng.2020.04.010.

[17] Yoo, Seung Hoon, et al. “Deep Learning-Based Decision-Tree Classifier for COVID-19 Diagnosis From Chest X-Ray Imaging.” Frontiers in Medicine, vol. 7, 2020, doi:10.3389/fmed.2020.00427.

[18] Hemdan, Ezz El-Din, et al. “COVIDX-Net: A Framework of Deep Learning Classifiers to Diagnose COVID-19 in X-Ray Images.” 24 March 2020, arXiv:2003.11055.

[19] Wang, Linda, et al. “COVID-Net: A Tailored Deep Convolutional Neural Network Design for Detection of COVID-19 Cases from Chest X-Ray Images.” 22 March 2020, arXiv:2003.09871.

[20] Kermany, Daniel; Zhang, Kang; Goldbaum, Michael (2018), “Labeled Optical Coherence Tomography (OCT) and Chest X-Ray Images for Classification”, Mendeley Data, v2, doi: 10.17632/rscbjbr9sj.2

[21] Kermany, Daniel S., et al. “Identifying Medical Diagnoses and Treatable Diseases by Image-Based Deep Learning.” Cell, vol. 172, no. 5, 2018, doi:10.1016/j.cell.2018.02.010.

[22] Jaeger, Stefan, et al. “Two Public Chest X-Ray Datasets for Computer-Aided Screening of Pulmonary Diseases.” Quantitative Imaging in Medicine and Surgery, 4 Dec. 2014, dx.doi.org/10.3978%2Fj.issn.2223-4292.2014.11.20.

[23] M.E.H. Chowdhury, T. Rahman, A. Khandakar, R. Mazhar, M.A. Kadir, Z.B. Mahbub, K.R. Islam, M.S. Khan, A. Iqbal, N. Al-Emadi, M.B.I. Reaz, M. T. Islam, “Can AI help in screening Viral and COVID-19 pneumonia?” IEEE Access, Vol. 8, 2020, pp. 132665-132676.

[24] Singh, Tarandeep. “COVID-19 & Normal Posteroanterior(PA) X-Rays.” Kaggle, 13 May 2020, www.kaggle.com/tarandeep97/covid19-normal-posteroanteriorpa-xrays.

[25] Patel, Prashant. “Chest X-Ray (Covid-19 & Pneumonia).” Kaggle, 18 June 2020, www.kaggle.com/prashant268/chest-xray-covid19-pneumonia.

[26] Raddar. “Chest X-Rays Tuberculosis from India.” Kaggle, Jaypee University of Information Technology, 19 Feb. 2020, www.kaggle.com/raddar/chest-xrays-tuberculosis-from-india.

[27] Larxel. “COVID-19 X Rays.” Kaggle, 18 Mar. 2020, www.kaggle.com/andrewmvd/convid19-x-rays.

[28] Huang, Gao, et al. “Densely Connected Convolutional Networks.” *ArXiv,* 25 Aug. 2016, pp. 1-1., arxiv.org/abs/1608.06993.

[29] Uvaradweb, and UVA | Physician Resource. “COVID-19 and Imaging: Why CT Scans and X-Rays Are Not Recommended for Diagnosing Coronavirus” UVA Radiology and Medical Imaging Blog for Patients, 21 Aug. 2020, blog.radiology.virginia.edu/covid-19-and-imaging/.

